# SCIseg: Automatic Segmentation of T2-weighted Intramedullary Lesions in Spinal Cord Injury

**DOI:** 10.1101/2024.01.03.24300794

**Authors:** Enamundram Naga Karthik, Jan Valosek, Andrew C. Smith, Dario Pfyffer, Simon Schading-Sassenhausen, Lynn Farner, Kenneth A. Weber, Patrick Freund, Julien Cohen-Adad

## Abstract

**Purpose:** To develop a deep learning tool for the automatic segmentation of T2-weighted intramedullary lesions in spinal cord injury (SCI).

**Material and Methods:** This retrospective study included a cohort of SCI patients from three sites enrolled between July 2002 and February 2023. A deep learning model, `SCIseg`, was trained in a three-phase process involving active learning for the automatic segmentation of intramedullary SCI lesions and the spinal cord. The data consisted of T2-weighted MRI acquired using different scanner manufacturers with heterogeneous image resolutions (isotropic/anisotropic), orientations (axial/sagittal), lesion etiologies (traumatic/ischemic/hemorrhagic) and lesions spread across the cervical, thoracic and lumbar spine. The segmentations from the proposed model were visually and quantitatively compared with other open-source baselines. Wilcoxon signed-rank test was used to compare quantitative MRI biomarkers (lesion volume, lesion length, and maximal axial damage ratio) computed from manual lesion masks and those obtained automatically with `SCIseg` predictions.

**Results:** MRI data from 191 SCI patients (mean age, 48.1 years ± 17.9 [SD]; 142 males) were used for model training and evaluation. SCIseg achieved the best segmentation performance for both the cord and lesions. There was no statistically significant difference between lesion length and maximal axial damage ratio computed from manually annotated lesions and those obtained using `SCIseg`.

**Conclusion:** Automatic segmentation of intramedullary lesions commonly seen in SCI replaces the tedious manual annotation process and enables the extraction of relevant lesion morphometrics in large cohorts. The proposed model segments lesions across different etiologies, scanner manufacturers, and heterogeneous image resolutions. `SCIseg` is open-source and accessible through the Spinal Cord Toolbox.

**Summary:** Automatic segmentation of the spinal cord and T2-weighted lesions in spinal cord injury on MRI scans across different treatment strategies, lesion etiologies, sites, scanner manufacturers, and heterogeneous image resolutions.

**Key Results:**

- An open-source, automatic method, `SCIseg`, was trained and evaluated on a dataset of 191 spinal cord injury patients from three sites for the segmentation of the spinal cord and T2-weighted lesions.
- `SCIseg` generalizes across traumatic and non-traumatic lesions, scanner manufacturers, and heterogeneous image resolutions, enabling the automatic extraction of lesion morphometrics in large multi-site cohorts.
- Quantitative MRI biomarkers, namely, lesion length and maximal axial damage ratio derived from the automatic predictions showed no statistically significant difference when compared with manual ground truth, implying reliability in `SCIseg’s` predictions.

## 1. Introduction

Spinal cord injury (SCI) refers to damage to the spinal cord (SC) due to traumatic or non-traumatic processes. Traumatic SCI results from acute damage to the spinal cord (SC) due to external physical factors (1,2). The majority of traumatic SCI patients sustain permanent neurological deficits such as motor and autonomic dysfunction with devastating physical and social consequences (1). Degenerative cervical myelopathy (DCM), the most common form of non-traumatic SCI, originates from chronic mechanical compression of the spinal cord (3). While relatively less common than traumatic lesions, ischemic SCI lesions represent up to 20% of all non-traumatic lesions (4,5) and show a similar course of recovery to traumatic SCI (6,7). MRI provides macrostructural information about the level of injury, intramedullary abnormalities (e.g., edema and hemorrhage), and allows the evaluation of soft tissue structures (1,3). Importantly, MRI-derived quantitative biomarkers, namely, intramedullary lesion length and lesion volume, have demonstrated associations with the neurological prognosis of traumatic SCI patients (7–12).

Despite recent advances in automatic SC MRI processing (13–16), robust methods for automatic quantitative MRI biomarker identification in SCI are still missing. As a result, most studies involve manual identification of these biomarkers (7,11,17–20), which is a time-consuming process potentially susceptible to inter-rater variability across sites, making it less reproducible in multi-site studies (3). Furthermore, segmentation of intramedullary SCI lesions in MRI scans poses an extremely challenging task mainly due to the evolving appearance of lesions in different injury phases (e.g., acute, sub-acute, intermediate) (1,2). The surgical implants in postoperative MRI scans might also cause severe image artifacts. Deep learning (DL) can improve the diagnosis and prognostication in SCI by automating the lesion annotation process, thereby reducing rater-specific biases and facilitating the analysis of large SCI cohorts across sites (21–23). Indeed, quantitative SCI lesion biomarkers derived from DL-based automatic segmentations have been shown to correlate well with clinical measures of motor impairment (24). However, despite its numerous advantages, DL has not been sufficiently explored in the context of SCI (22), with no open-source methods existing to date. This suggests a need for an automatic biomarker identification method that deals with the complex pathophysiology of SCI patients, generalizes to multiple sites and is easily accessible by researchers.

Our objective was to develop an open-source DL-based tool, *`SCIseg`*, for the automatic segmentation of the spinal cord and intramedullary lesions from T2-weighted MRI scans of SCI patients [R1.10]. We evaluated two hypotheses: first, that quantitative MRI biomarkers derived automatically, such as lesion volume, lesion length, and maximal axial damage ratio, would not significantly differ from those identified manually. Second, we hypothesized that these biomarkers would correlate with clinical measures post-SCI. To this end, we conducted correlation analyses between biomarkers derived using the automatic `SCIseg` and clinical scores, specifically pinprick, light touch, and lower extremity motor scores. [R1.9, R2.1]

## 2. Materials and Methods

### 2.1 Study Design and Participants

This retrospective study included a cohort of 191 SCI patients from three sites enrolled between July 2002 and February 2023. All patients provided written informed consent following Institutional Review Board approval and the Declaration of Helsinki. From site 1, 97 patients were enrolled out of which 61 had surgical hardware (dorsal/ventral spondylodesis), 13 underwent decompressive surgery, and the remaining 23 patients did not undergo surgery. From site 2, 80 patients were enrolled, all of which had post-operative metallic stabilization. Lastly, 14 patients were enrolled from site 3, out of which 8 had surgical hardware, 2 had decompressive surgery and 4 patients did not undergo any surgery. Details on patient demographics, injury levels, injury chronicity, scanner types, etc. can be found in Table 1. [AC, DE]. The inclusion criteria were: traumatic or ischemic SCI, patients with and without hardware, clinical MRI available for analyses, and completed enrollment. Exclusion criteria were: concurrent traumatic brain injury beyond concussion, and significant pre-existing neurological history (i.e., multiple sclerosis, transverse myelitis, cerebrovascular stroke). Patients from site 2 were clinically assessed using the international standards for the neurological classification of SCI (ISNCSCI) protocol (25) to obtain light touch, pinprick, and lower extremity motor scores as previously described in studies by Smith et al. (11,26). Patients from all sites were reported previously (7,11,17,26,27). These articles used manually annotated lesion masks to study the clinical consequences of SCI and their predictive relationships with motor and sensory functions. In contrast, our study presents a DL-based tool to automatically segment intramedullary SCI lesions.

**Table 1:** Characteristics of the Study Cohort.

|  | Site 1 | Site 2 | Site 3 |
| --- | --- | --- | --- |
| **Number of patients** | 97 | 80 | 14 |
| **Number of MRI scans** | 137[R1.29]<sup>†</sup> | 80 | 14 |
| **Sex**<br>**(male/female)** | 66/25* | 65/15 | 11/2 |
| **Age**<br>**(mean ± standard deviation)** | 51.0 ± 19.1 | 45.8 ± 16.4 | 42.9 ± 16.7 |
| **Age range** | 17–83 | 15–81 | 21–65 |
| **Days from injury to MRI**<br>**(mean ± standard deviation)** | 376.2 ± 1364.4<sup>‡</sup> | 84.0 ± 212.1 | 579.1 ± 714.1 |
| **Days from injury to MRI**<br>**(median)** | 41.5 | 21.0 | 407 |
| **Number of patients**<br>**with surgical**<br>**implants/hardware** | 61<br>3T (n=19), 1.5T (n=41), 1T (n=1) | 80<br>3T (n=21), 1.5T (n=59) | 8<br>3T (n=10) |
| **Lesion origin** [R1.13, R1.14, R3.4] | Traumatic (n=77)<br>Ischemic (n=13)<br>Hemorrhagic (n=5)<br>Unknown origin (n=3) | Traumatic (n=80) | Traumatic (n=14) |
| **Injury level** [R3.2, R3.6] | Cervical (n=58)<br>Thoracic (n=31)<br>Lumbar (n=8) | Cervical (n=80) | Cervical (n=14) |
| **Number of patients in**<br>**train set** | Cervical (n=44)<br>Thoracic (n=28)<br>Lumbar (n=6)<br>*Total (n=78)* | Cervical (n=64) | Cervical (n=14) |
| **Number of patients in**<br>**test set** | Cervical (n=14)<br>Thoracic (n=3)<br>Lumbar (n=2)<br>*Total (n=19)* | Cervical (n=16) | 0 |
| **MRI manufacturers** | Siemens (n=91), GE (n=5), Philips (n=1) | Siemens (n=20), GE (n=60) | Siemens (n=14) |
| **MRI field strength** | 3T (n=37), 1.5T (n=59), 1T (n=1) | 3T (n=21), 1.5T (n=59) | 3T (n=14) |
| **MRI Sequence parameters** | SAGITTAL T2-weighted: voxel size $0.34 \times 0.34$ mm to $0.96 \times 0.96$ mm; slice thickness 2.2 mm to 4.8 mm | AXIAL T2-weighted: voxel size $0.31 \times 0.31$ mm to $0.78 \times 0.78$ mm; slice thickness from 3.0 mm to 6.0 mm | ISOTROPIC T2-weighted: voxel size $0.84 \times 0.84 \times 0.94$ mm to $0.875 \times 0.875 \times 0.9$ mm |
| | AXIAL T2-weighted: voxel size $0.35 \times 0.35$ mm to $0.78 \times 0.78$ mm; slice thickness 1.0 mm to 7.0 mm | | |

\*Sex not reported for 7 patients
†Eight patients were followed up with more than 1 MRI examination
‡Five scans were acquired very late after injury resulting in high average time for MRI examination

### 2.2 MRI Data

The MRI scans were converted from DICOM to NIfTI format and organized according to the BIDS standard (28) at individual sites. During this curation process, the scans were anonymized (i.e., all sensitive patient information was deleted). T2-weighted (T2w) MRI scans with varying lesion etiologies (traumatic, ischemic, hemorrhagic), injury chronicity (sub-acute, intermediate, and chronic), orientations (sagittal/axial), and voxel sizes were used for this study (Figure 1, Table 1). Lesions appearing as T2w signal abnormalities (hyperintense or hypointense voxels corresponding to primary contusions, secondary cytotoxic edema or hemorrhage [R1.13, R1.14]) were manually annotated as a single object by two raters from site 1, one rater from site 2, and one rater from site 3 using JIM and FSLeyes image viewers. As obtaining the ground truth (GT) SC segmentation masks using a fully manual approach is time-consuming, `sct_deepseg_sc` (29) was used to initially segment the SC for all 3 sites, followed by manual corrections wherever necessary. Such semi-automatic approaches were also reported in previous studies (29,30). [R1.21, R2.6]

**Figure 1:**
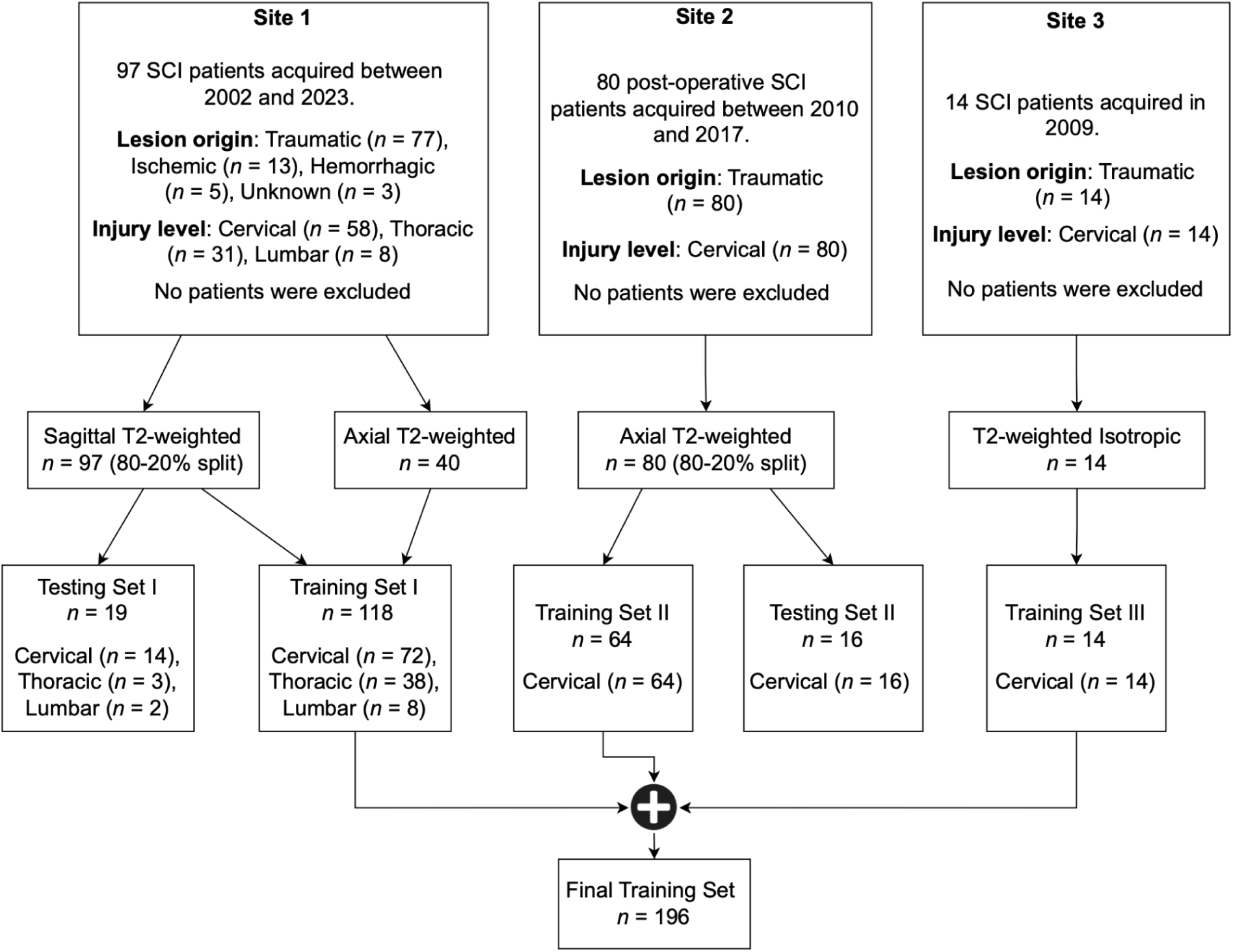
Study Flowchart. The data included patient cohorts from three sites with heterogeneous image resolutions, orientations, and lesion etiologies. The validation set is included within the final training set. Models were evaluated independently on the test sets of Site 1 and Site 2. Please refer to Table 1 for details on the MRI vendors and field strengths.

### 2.3 Deep Learning Training Protocol

The model was trained in three phases (Figure 2). In the initial phase, a baseline segmentation model was trained using a labelled dataset of 78 subjects with T2-weighted sagittal scans (site 1) and 64 subjects with T2-weighted axial scans (site 2). We used the *region-based training* strategy of nnUNet (31), where the model initially segments the SC and then localizes itself on the SC to segment the T2-weighted lesions subsequently. The lesions are segmented as a single object covering hyperintense and hypointense voxels hence containing both edema and hemorrhage. [R1.14, R1.25] Default data augmentation methods by nnUNet were used, namely, random rotation, scaling, mirroring, Gaussian noise addition, Gaussian blurring, adjusting image brightness and contrast, low-resolution simulation, and Gamma transformation. All scans were preprocessed with RPI orientation and Z-score normalization. The model was trained for 1000 epochs, with a batch size of 2 using the stochastic gradient descent optimizer with a polynomial learning rate scheduler.

**Figure 2:**
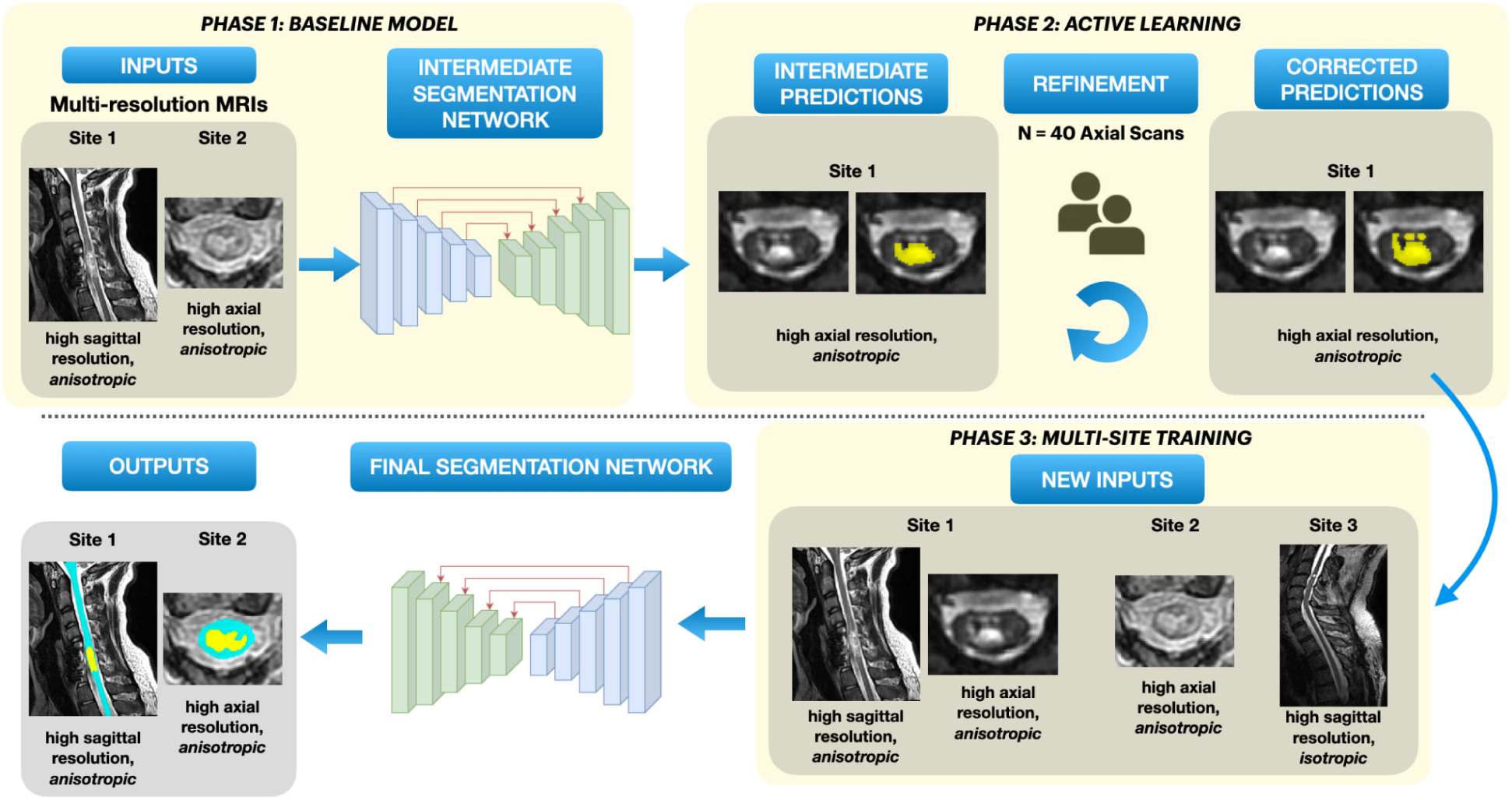
Overview of our segmentation approach. Phase 1: A baseline model is trained on data consisting of T2-weighted scans with axial and sagittal orientations from two sites. Phase 2: *Active learning* – Initial batch of automatic predictions on T2-weighted axial scans from site 2 are obtained, followed by manual corrections. Phase 3: Along with the newly corrected axial scans, isotropic T2-weighted sagittal scans from site 3 are added to the original dataset for multi-site training. The final model is trained to segment both spinal cord and lesion simultaneously.

For the second phase, we used the human-in-the-loop active learning strategy (32) to gradually include axial T2-weighted scans from site 1 in the training dataset. Using the phase 1 baseline model, we generated initial SC and lesion predictions for unlabeled axial scans from site 1. A subset of predicted segmentations underwent quality control, with two raters manually correcting if needed. These refined segmentations were then added to the training dataset, resulting in the inclusion of 40 scans and leading to a total of 182 scans in the training set.

To further improve our model’s generalization capabilities to a wide range of image resolutions, we added a new dataset from site 3 containing 14 *isotropic* T2-weighted sagittal scans of traumatic SCI patients in the third training phase. In summary, the final dataset consisting of 196 scans gathered from three sites was used for training the model with the region-based strategy described above.

### 2.4 Evaluation Protocol

We created two independent test sets (site 1: n=19, site 2: n=16), following the 80/20 train/test splitting ratio (Table 1). To ensure an unbiased assessment of the model’s performance and avoid overfitting, the train/test splits were done at the patient-level (and not at the image-level), ensuring that sagittal and axial scans of a particular patient strictly belonged only to the training or testing set. [R1.12, R1.16] We trained five models, each with a different train/test splitting using a different random seed, to avoid biasing the model towards a particular dataset split. The model’s performance on the lesion and SC segmentation were evaluated independently within each test set by comparing it with open-source methods available in Spinal Cord Toolbox (SCT) (15): `sct_propseg` (33)`, sct_deepseg_sc` (29), and the recently proposed `contrast-agnostic` SC segmentation model (30). Due to the lack of existing state-of-the-art, open-source methods for SCI lesion segmentation, we compared the `SCIseg` 3D model with its 2D version. Additionally, we tested our model on an independent cohort of 14 DCM patients from site 1 (unseen during training) to evaluate its generalization on non-traumatic SCI patients.

### 2.5 Evaluation Metrics

For quantitative validation, we used the segmentation metrics from the open-source ANIMA toolkit (https://anima.readthedocs.io/en/latest/index.html). For SC segmentation, we presented the Dice coefficient and the relative volume error (RVE). For lesion segmentation, we reported the Dice coefficient, average surface distance, lesion-wise positive predictive value (PPV), lesion-wise sensitivity, and F_1_ score (34). As we trained five models on five random train/test splits, some patients were present in more than one test set. We thus averaged the metrics across test splits for such patients.

### 2.6 Quantitative MRI Biomarkers

We used the SCT’s `sct_analyze_lesion` function to automatically compute the total lesion volume, intramedullary lesion length, and maximal axial damage ratio (11) from the manual ground-truth lesion masks and the automatic predictions using the proposed `SCIseg` 3D model. To assess the effect of adding more training data during active learning, we computed the quantitative MRI biomarkers before (phase 1) and after active learning (phase 3). The quantitative MRI biomarkers were then averaged across five random test splits. Additionally, for site 2, we correlated the quantitative MRI biomarkers with the clinical scores (light touch, pinprick, and lower extremity motor scores).

### 2.7 Statistical Analysis

Statistical analysis was performed using the SciPy Python library v1.11.4 (35). Data normality was tested using D’Agostino and Pearson’s normality test. Between-group comparisons (lesion segmentation performance `SCIseg` 2D vs. `SCIseg` 3D; `SCIseg` lesion segmentation performance before (phase 1) vs. after active learning (phase 3); manual lesion GT vs. `SCIseg`-predicted lesions) were performed using the Wilcoxon signed-rank test. Correlations between clinical scores and quantitative MRI biomarkers were examined using the Spearman rank-order correlation.

## 3. Results

### 3.1 Patient Characteristics

A total of 191 patients (mean age ± standard deviation 48.1 ± 17.9, 142 males, 42 females, 7 sex not reported) with 231 MRI scans from three sites with different lesion etiologies (traumatic/ischemic/hemorrhagic) were included in this study (Figure 1, Table 1). Eight patients from site 1 were followed up with additional MRI examinations. Patients were scanned across scanners from different manufacturers (Siemens, Philips, GE) with different field strengths (1T, 1.5T, 3T). T2-weighted scans used in this study had heterogeneous image resolutions, and orientations (Table 1).

### 3.2 Automatic Spinal Cord and Lesion Segmentation in SCI

Table 2 shows the quantitative results of `SCIseg 3D` on test sets of the two sites. We observed that SC segmentations from the model are quite stable across different data splits despite the presence of artifacts in the scans. However, for lesion segmentation, the model showed better performance on site 2 compared to site 1 with a high standard deviation across splits.

**Table 2.**
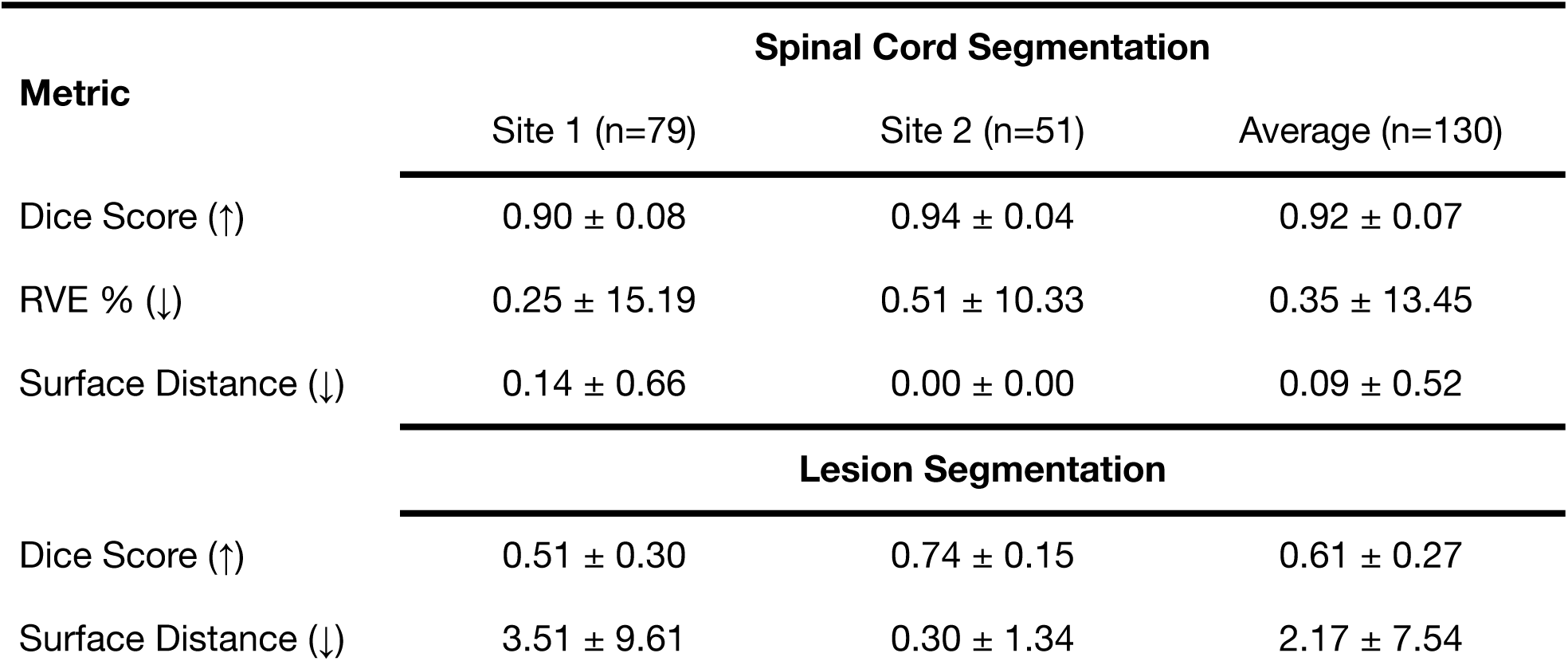
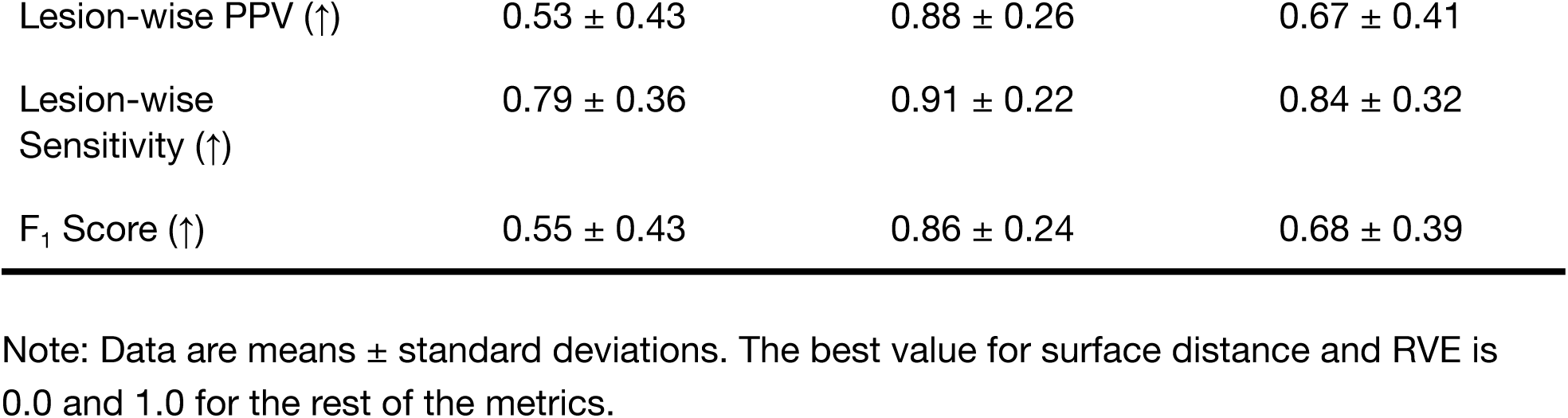
Quantitative performance of the proposed SCIseg 3D model. The metrics are averaged across 5 random seeds.

### 3.3 Comparison with Other Methods

We compared the SC segmentation performance of our `SCIseg 3D` model with other methods: `sct_propseg`, `sct_deepseg_sc 2D`, `sct_deepseg_sc 3D`, `contrast-agnostic`, and `SCIseg 2D` (Figure 3, Figure 4). The half-violin plots in Figure 4 show the distribution of the Dice scores and RVE for test scans across all seeds and the scatter plots show the performance of the models on each test scan.

**Figure 3:**
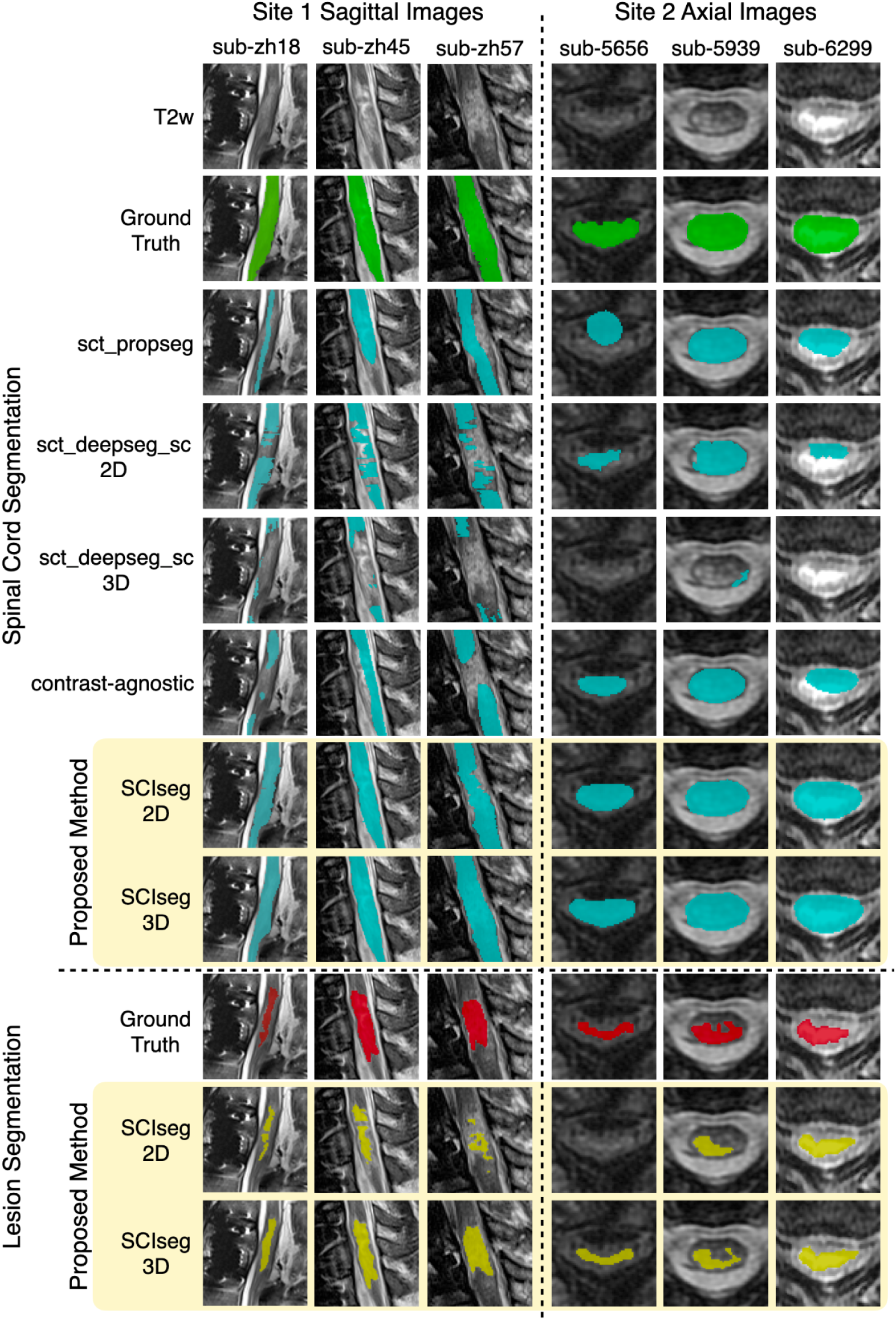
Comparison of `SCIseg` with baseline methods for the spinal cord and lesion segmentation on patients from site 1 and site 2. Notice that `SCIseg` 3D provides the best results qualitatively for both spinal cord and lesion segmentation at the site of lesions.

**Figure 4:**
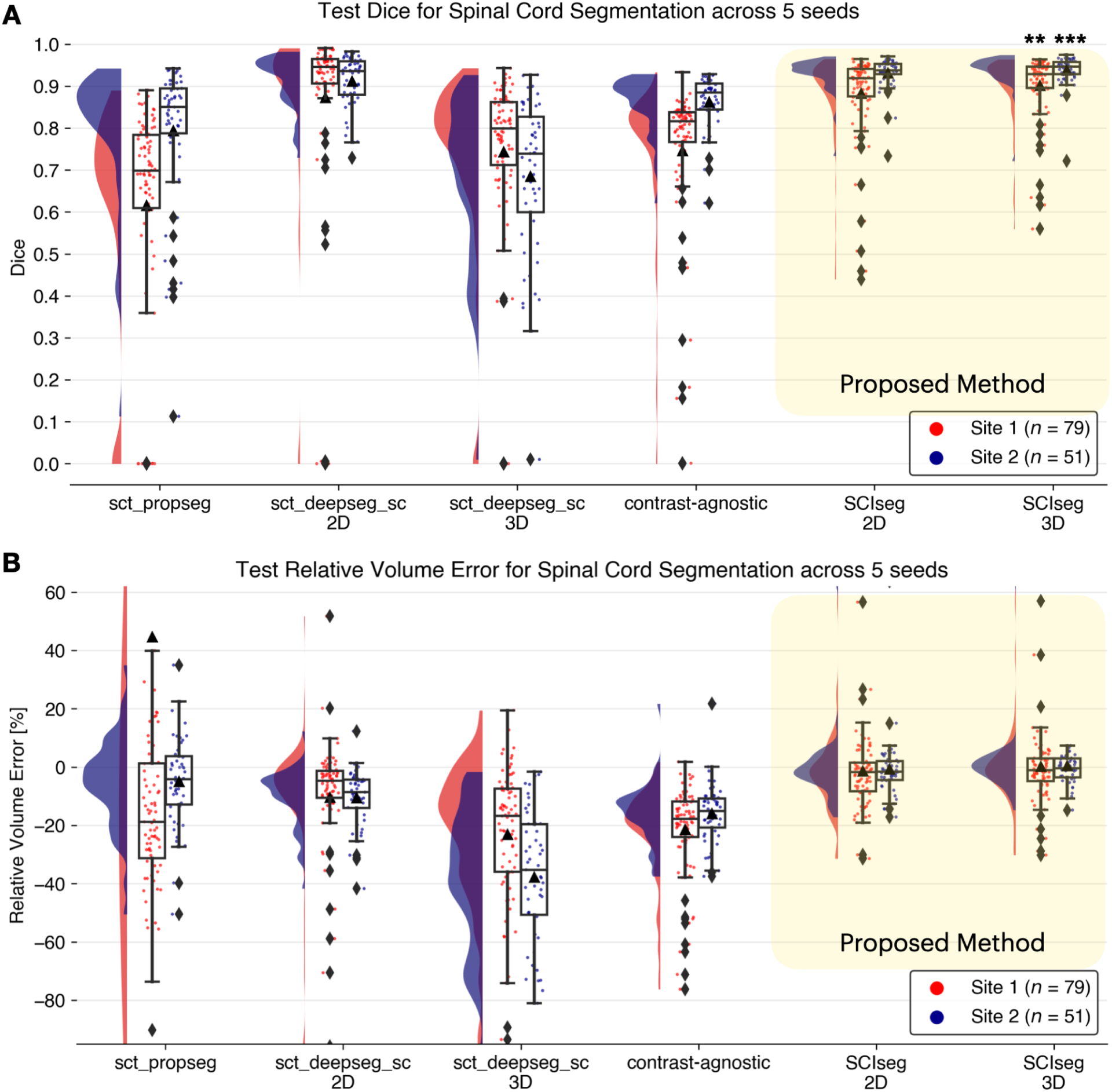
Raincloud plots comparing the (A) Dice scores (best: 1; worst: 0) and (B) relative volume error (in %, best: 0%) across various spinal cord segmentation methods. The numbers in the legend represent the number of test scans in each site across 5 different training seeds. Notice that although the `sct_deepseg_sc` 2D and `SCIseg` 3D have similar Dice scores, the former shows a higher under-segmentation (negative relative volume error) compared to the latter. *** *P* < .05 (two-sided Bonferroni-corrected pairwise Wilcoxon signed-rank test for `SCIseg` 3D with all baselines), ** *P* < .001 (statistically significant for all pairs except `SCIseg` 3D and `sct_deepseg_sc` 2D).

#### 3.3.1 Spinal Cord Segmentation

Our model, `SCIseg 3D`, achieves the best segmentation performance across all baselines (Figure 4). For site 1, `SCIseg 3D` model outputs SC segmentations for all test cases including those where the baselines output empty predictions (shown by diamonds at Dice=0 in Figure 4A). We also observed more under-/over-segmented predictions for site 1 (shown by a larger spread of scatter points around RVE=0% in Figure 4B. On visual quality control of such cases, we noticed that the scans contained substantial metal implants, interfering with our model’s ability to fully segment the SC. On the other hand, for site 2, our model performed robustly across all test scans. Figure 4A shows that the distribution of Dice scores for one of the baseline models (`sct_deepseg_sc 2D`) is similar to `SCIseg 3D`. As described in Section 2.2, this is the consequence of the fact that the semi-automatic approach involving `sct_deepseg_sc 2D` was used to create the GT SC masks. As a result, quantitative evaluations involving the GT masks obtained from this baseline model are inherently biased to be higher than the rest of the methods in comparison. Lastly, it must be noted that all baselines were trained specifically for segmenting the spinal cord, whereas the proposed `SCIseg 3D` model can segment both SC and lesions simultaneously.

#### 3.3.2 Lesion Segmentation

Table 3 presents a comparison between the 2D and 3D variants of the `SCIseg` model. The 3D model performs significantly better than the 2D model across all metrics for both sites. As for the performance within sites, the model’s performance on site 2 is higher than that of site 1. Through visual quality control, we noticed that site 1 contained several patients with metal implants causing heavy image artifacts and patients spanned different SCI phases (acute and sub-acute) with various degrees of lesion hyperintensity, thus making automatic segmentation challenging. Despite these issues, the `SCIseg` model provides a good starting point for obtaining lesion segmentations instead of manually annotating lesions from scratch.

**Table 3.**
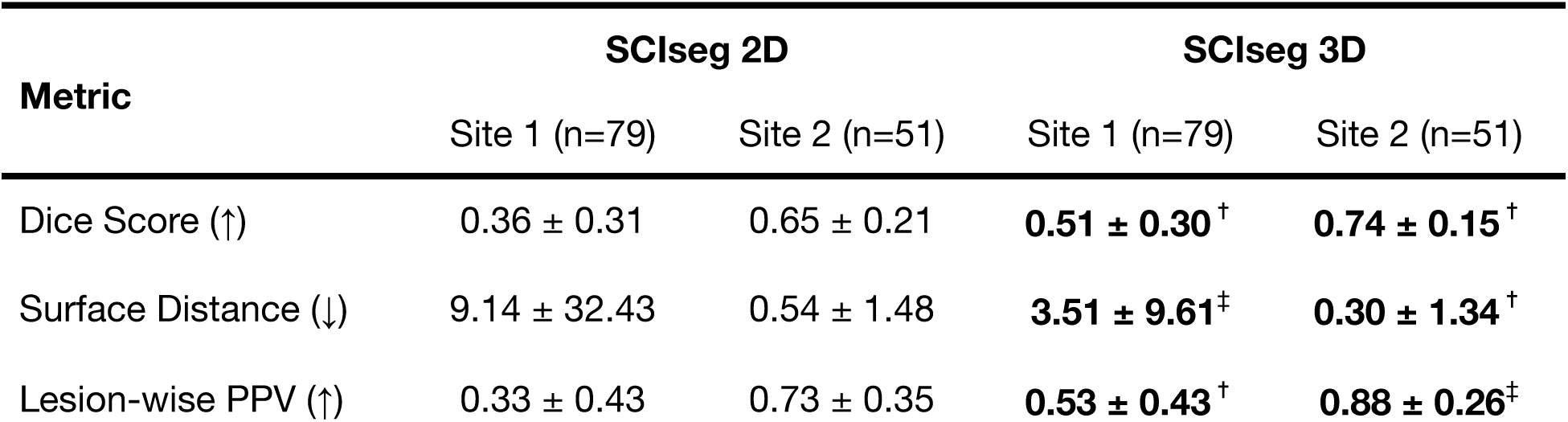
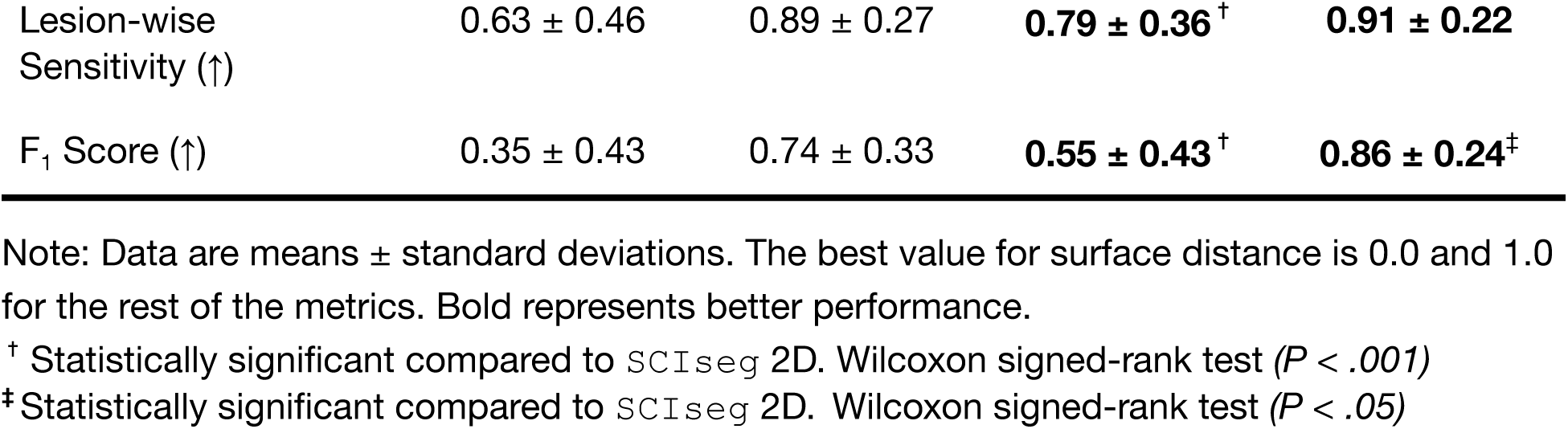
Lesion segmentation performance of the SCIseg models. The metrics are averaged across 5 different training seeds.

| Metric | SCIseg 2D |  | SCIseg 3D |  |
| --- | --- | --- | --- | --- |
|  | Site 1 (n=79) | Site 2 (n=51) | Site 1 (n=79) | Site 2 (n=51) |
| Dice Score (↑) | 0.36 ± 0.31 | 0.65 ± 0.21 | **0.51 ± 0.30<sup>†</sup>** | **0.74 ± 0.15<sup>†</sup>** |
| Surface Distance (↓) | 9.14 ± 32.43 | 0.54 ± 1.48 | **3.51 ± 9.61<sup>‡</sup>** | **0.30 ± 1.34<sup>†</sup>** |
| Lesion-wise PPV (↑) | 0.33 ± 0.43 | 0.73 ± 0.35 | **0.53 ± 0.43<sup>†</sup>** | **0.88 ± 0.26<sup>‡</sup>** |
| Lesion-wise<br>Sensitivity (↑) | $0.63 \pm 0.46$ | $0.89 \pm 0.27$ | **$0.79 \pm 0.36^{\dagger}$** | **$0.91 \pm 0.22$** |
| $F_1$ Score (↑) | $0.35 \pm 0.43$ | $0.74 \pm 0.33$ | **$0.55 \pm 0.43^{\dagger}$** | **$0.86 \pm 0.24^{\ddagger}$** |

Note: Data are means ± standard deviations. The best value for surface distance is 0.0 and 1.0 for the rest of the metrics. Bold represents better performance.
$^{\dagger}$ Statistically significant compared to *SCIseg* 2D. Wilcoxon signed-rank test ( $P < .001$ )
$^{\ddagger}$ Statistically significant compared to *SCIseg* 2D. Wilcoxon signed-rank test ( $P < .05$ )

### 3.4 Effect of Active Learning on Lesion Segmentation

We performed an ablation study comparing the model performance after phase 1 (training on 2 sites) and phase 3 (training on 3 sites after active learning). Figure 5A shows the correlation between manual ground truth and automatic predictions for total lesion volume (top) and intramedullary lesion length (bottom). For both sites, a higher agreement between the manually annotated and automatically derived lesion metrics can be observed for the final model after the third phase of training (i.e., solid lines moving closer to the diagonal identity line). The improvement after active learning (phase 3) was statistically significant (Wilcoxon signed-rank test, p < 0.05) in estimating the total lesion volume for both sites and only the lesion length for site 1.

**Figure 5:**
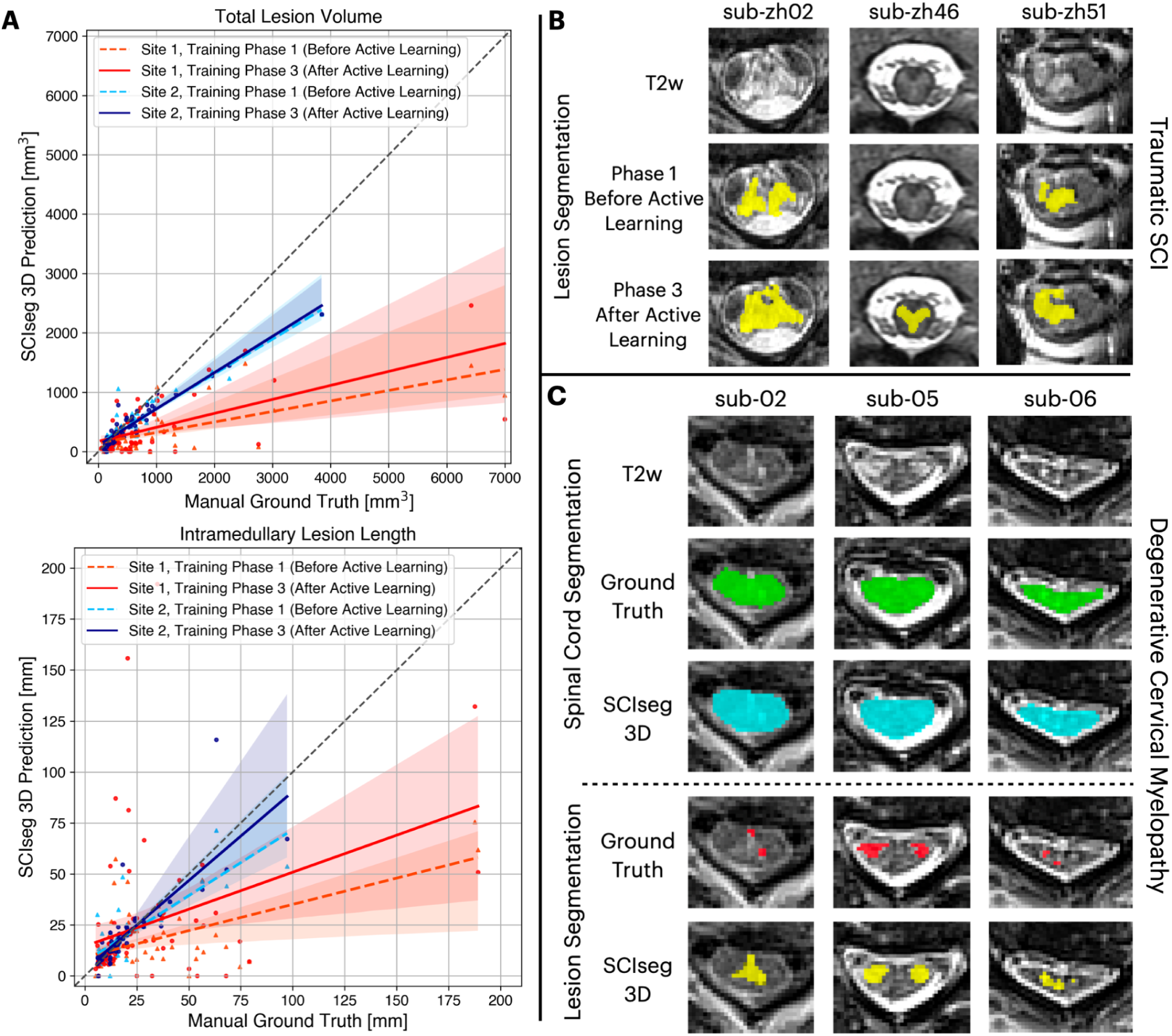
Comparison of model performance before and after active learning. (A) Correlation plots for total lesion volume (top) and intramedullary lesion length (bottom) computed from manual ground truth (GT) lesion masks (x-axis) and lesion predictions from the proposed `SCIseg` 3D model (y-axis). Within each plot, coloured dashed and solid lines show the agreement between manual GT and automatic predictions before and after active learning, respectively, site 1 (red/orange) and site 2 (blue/light-blue). Note that the model’s predictions after active learning show a higher agreement with the manual GT for both sites (i.e., solid lines move closer to the diagonal identity line). (B) SCIseg’s predictions on unseen axial images from site 2 before and after active learning. (C) Examples of SCIseg’s generalization to non-traumatic SCI (i.e., degenerative cervical myelopathy, DCM) patients. Notice that the model obtains an accurate SC segmentation even at the level of severe compression (sub-06).

Figure 5B shows the performance of our baseline model after phase 1 of training (before active learning) on unseen axial T2-weighted scans from site 1. The model tends to under-segment the lesions. However, we noticed an overall improvement in the segmentations when trained on more data consisting of axial scans from site 1 and isotropic sagittal scans from site 3 during phase 3 of training.

### 3.5 Generalization to Degenerative Cervical Myelopathy

Qualitative examples of SC and lesion segmentation on an independent cohort of DCM patients unseen during training are shown in Figure 5C. Interestingly, in cases where the ground-truth lesion masks were under-segmented, the model provided a better and more complete segmentation of the lesion. Furthermore, the SC segmentations are accurate even for slices with severe SC compression (Figure 5C, `sub-06`). For quantitative validation, we also computed the Dice and F_1_ scores for both SC and lesion segmentations (Table 4).

**Table 4.** Quantitative evaluation of SCIseg 3D model’s generalization to non-traumatic SCI (i.e., DCM) patients. . The metrics are averaged across 5 random seeds.

| Metric | Spinal Cord Segmentation |
| --- | --- |
| Dice Score (↑) | $0.95 \pm 0.01$ |
| RVE % (↓) | $3.51 \pm 4.63$ |
| Surface Distance (↓) | $0.00 \pm 0.00$ |
|  | Lesion Segmentation |
| Dice Score (↑) | $0.46 \pm 0.26$ |
| Surface Distance (↓) | $1.51 \pm 4.64$ |
| Lesion-wise PPV (↑) | $0.71 \pm 0.38$ |
| Lesion-wise Sensitivity (↑) | $0.50 \pm 0.45$ |
| $F_1$ Score (↑) | $0.49 \pm 0.44$ |

Note: Data are means ± standard deviations. The best value for surface distance and RVE is 0.0 and 1.0 for the rest of the metrics.

### 3.6 Correlation between Clinical Scores and MRI Biomarkers

Figure 6 illustrates the relationship between clinical scores (specifically, pinprick, light touch, and lower extremity motor scores) and quantitative MRI biomarkers (namely, lesion volume, lesion length, and maximal axial damage ratio) calculated from both manual ground-truth lesion masks and lesions predicted using `SCIseg` 3D. We observed statistically significant correlations (P < .05) between the clinical scores and lesion biomarkers (Figure 6). The Wilcoxon signed-rank test between manual (yellow) vs. `SCIseg`-predicted (green) lesion biomarkers revealed no statistically significant (*P* > .05) difference for lesion length and maximal axial damage ratio, while lesion volume showed a significant difference (P < .05).

**Figure 6:**
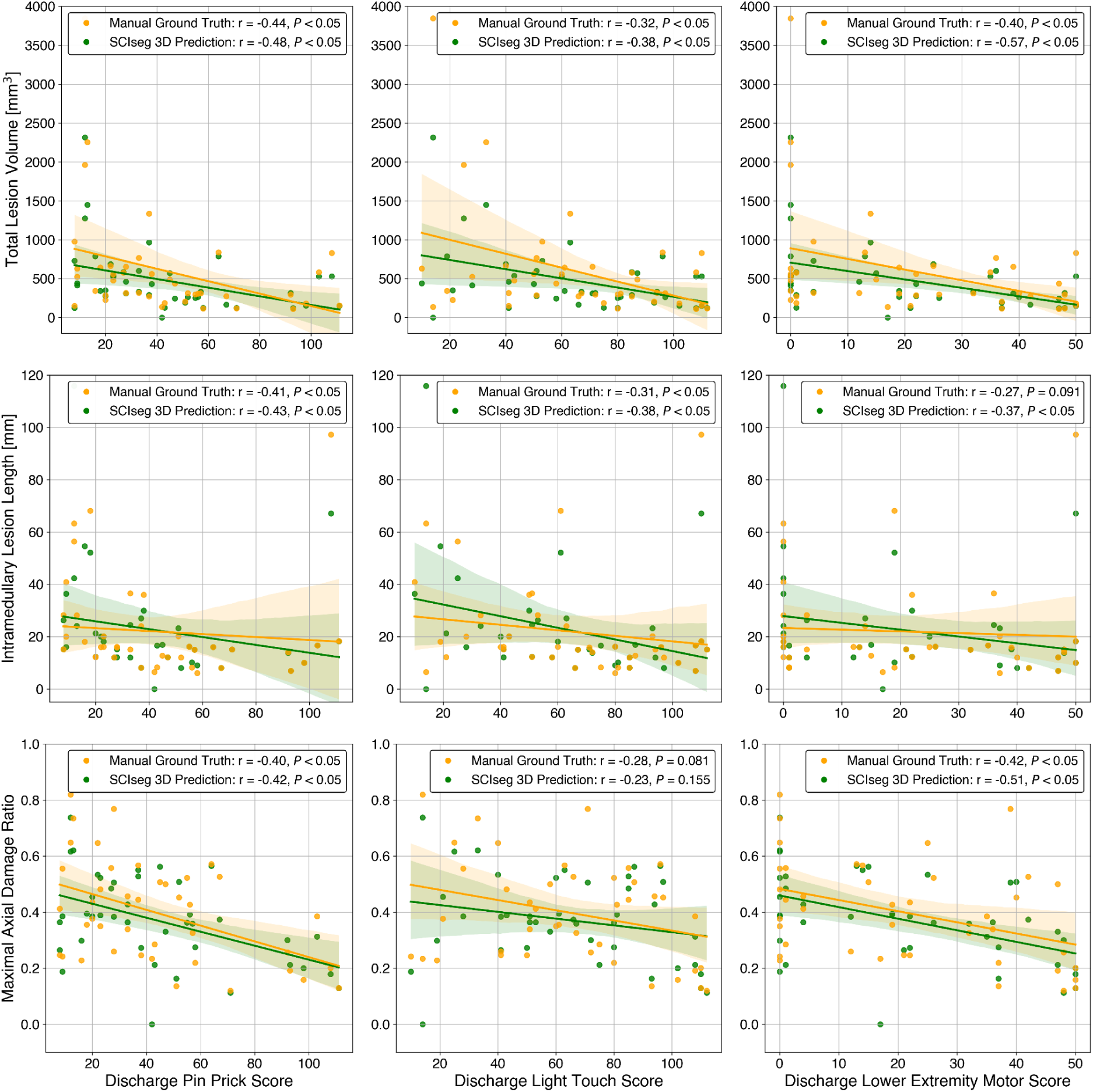
Correlation analysis between discharge clinical scores (x-axis) and quantitative MRI biomarkers (y-axis) for site 2. Spearman correlation coefficient and p-value are shown in the legends. The Wilcoxon signed-rank test between manual ground-truth lesion masks (yellow) vs. automatic predictions using SCIseg 3D (green) lesion biomarkers revealed no statistically significant (P > .05) difference for lesion length and maximal axial damage ratio, while lesion volume showed a significant difference (P < .05).

## 4. Discussion

This study introduced a DL-based model, `SCIseg`, for the automatic segmentation of the spinal cord and intramedullary lesions in SCI patients from T2-weighted MRI scans. The model was trained and evaluated on a cohort of 191 traumatic and non-traumatic SCI patients with 231 scans acquired using different scanner manufacturers with heterogeneous image resolutions (isotropic/anisotropic), orientations (axial/sagittal), lesion etiologies (traumatic/ischemic/hemorrhagic) and lesions spread across the cervical, thoracic and lumbar spine. To the best of our knowledge, `SCIseg` is the first open-source, automatic method for lesion and spinal cord segmentation in SCI. It also generalizes to DCM patients, producing accurate segmentations for both lesions and SC at the compression levels.

As the segmentation performance might be constrained by the low data quality and small dataset sizes in SCI, we showed that implementing a three-phase training strategy, including an active learning approach to progressively expand the dataset size and incorporating diverse data distributions into the training set, contributes significantly towards enhancing the model performance. Furthermore, a region-based training strategy that jointly segments the SC and the lesion is more efficient than training two individual models for SC and lesion segmentation, respectively. As a result, correlation analyses between clinical scores and MRI-derived biomarkers showed statistically significant relationships (*P* < .05) for both manually annotated ground truth and automatically derived lesion masks, suggesting the `SCIseg` predictions can be reliably used for correlation with clinical measures in SCI.

Our cohort predominantly consisted of traumatic SCI lesions in intermediate and chronic phases as the prevalence of ischemic and hemorrhagic lesions is typically lower (4). As chronic injuries tend to be more delineated on T2w scans (2), our model learned to be sensitive to hyperintense abnormalities in the image. This also explains its ability to segment DCM lesions which also tend to be hyperintense at the site of compression. Similarly, as the injury levels in the training dataset were skewed towards the cervical spine, its ability to segment lumbar lesions is expected to be lower compared to cervical/thoracic lesions. [R3.2, R3.8]

Only a few studies exist in the literature discussing the importance of automatic segmentation in SCI scans (24,36). McCoy et al.’s study (24) is the closest to ours as it presented the first DL method for the segmentation of SC and intramedullary lesions in SCI. Nevertheless, there are several important distinctions between the two studies. While their model was trained on axial scans of pre-operative (acute) SCI patients from a single site, our model was trained on multi-site data consisting of traumatic and ischemic SCI patients with different image orientations (axial/sagittal). Moreover, our model was exposed to more heterogeneous data covering different injury chronicities (intermediate/chronic) and therefore demonstrated better generalization to both traumatic and non-traumatic lesion etiologies. More importantly, our work is open-source, further enabling reproducible, multi-site studies in SCI.

This study has a few limitations. First, longitudinal scans from patients with follow-up examinations were treated as independent inputs for training. While the lesion appearance evolved between sessions, resulting in non-identical lesions (hence justifying our choice of treating them as independent inputs), the model was unlikely to learn the evolution of lesions across time. Second, the model’s sensitivity to hyperintense abnormalities might result in false positive segmentations in healthy controls where the SC central canal is visualized. Third, our limited training set size of 196 scans risks overfitting, given the complexity of the SCI lesion segmentation task. While we gathered diverse data from 3 sites and trained 5 models with different train/test splits along with extensive data augmentation to prevent potential overfitting, increasing the dataset size would further improve the model’s performance and generalization. Lastly, we did not analyze the inter-rater variability as the data were gathered from multiple sites and there were no overlapping subjects across sites. Previous studies (37,38) have reported that MRI measures of spinal cord damage (e.g., edema length, midsagittal tissue bridge ratio, axial damage ratio) exhibit high-to-excellent levels of inter-rater reliability.

There exist several promising avenues for future work. The segmentation models can be improved by using more fine-grained ground-truth masks, where the hyperintense edema and hypointense hemorrhage could be treated as separate classes. Training a model on pre-operative traumatic SCI data using these ground-truth masks would have a major impact on improving the initial classification of the disease and further prognostication (18). While the model generalizes reasonably well to DCM lesions, there is a scope for improvement, especially, by adding the DCM cohort to the existing training set or by training a DL model exclusively on DCM data. Previous studies have reported the presence of hyperintense T2-weighted lesions in up to 64% of DCM patients (13,39,40) and explored the relationship between structural and functional damages (41). Such studies would greatly benefit from an automatic DCM lesion segmentation method.

In conclusion, this study presented `SCIseg`, an automatic DL-based method for the segmentation of SC and intramedullary lesions in SCI from T2-weighted MRI scans. The work has addressed several limitations of previous studies, first, a large retrospective cohort consisting of 191 patients spanning three sites was used, second, MRI data was acquired using scanners from different manufacturers, and third, a single model was trained to segment *both* SC and lesions. More importantly, the methodology has been designed to ensure reproducibility and enable large-scale, reproducible prospective studies. The model is open-source and accessible via SCT (v6.2 and higher). We hope that `SCIseg` will benefit clinicians and patients by providing additional diagnostic and prognostic information, serving as a basis for further studies assessing optimal rehabilitation from a customized patient-based perspective.

## Data Availability

All data produced in the present study are available upon reasonable request to the authors

https://github.com/ivadomed/model_seg_sci

## List of Abbreviations

DCM: degenerative cervical myelopathy
DL: deep learning
RVE: relative volume error
SC: spinal cord
SCI: spinal cord injury
SCT: Spinal Cord Toolbox

## Code Availability Statement

To facilitate reproducibility and open science principles, all codes, processing scripts, and results are shared as open-source and freely available to the whole community at https://github.com/ivadomed/model_seg_sci.

## Acknowledgements

We thank Nick Guenther and Mathieu Guay-Paquet for their assistance with the management of the datasets, Joshua Newton for his contributions in helping us implement the algorithm to SCT, Maxime Bouthillier for his help in correcting parts of the manuscript, Drs. Thierry Albert, Bertrand Baussart, Caroline Hugeron, Hugues Pascal Moussellard, Frédéric Petit and Marc-Antoine Rousseau for helping with patient recruitment in Paris, Dr. Serge Rossignol and the Multidisciplinary Team on Locomotor Rehabilitation (Regenerative Medicine and Nanomedicine, CIHR), and we thank all patients.

